# A randomized trial on the regular use of potent mouthwash in COVID-19 treatment

**DOI:** 10.1101/2020.11.27.20234997

**Authors:** Khalid Mukhtar, Suelen Qassim, Shaikha Ali Al Qahtani, Mohamed Ibn-Masud Danjuma, Mohamed Mohamedali, Housamaddeen Al Farhan, Mohammed F. Khudair, Abdel Rehim El Tayeh, Mohammed Al-Dosari, Mohamed Elhassan Babiker, Ahmed Hassib, Rumaisa Mohamed Elmustafa, Wesal Elhadary, Morwan Abdulkarim, Rajvir Singh, Muna Al.Maslamani

**Affiliations:** Hamad Medical Corporation, Doha, QATAR; Qatar University – Public Health program, Doha, Qatar

## Abstract

In this work we tried to study the effect of the regular use of potent mouthwash in COVID19 cases, on the premise that it may speedup the recovery, through the repeated reduction of microbial load, of both, the 2019-nCOV and oral microbiota; thus slowing the disease progression and lowering the incidence of superinfections.

Through a randomized controlled trial, a mixed solution of Hydrogen peroxide 2% and chlorhexidine gluconate, to be used for oral rinsing and gargling three times daily, was tested in cases admitted to COVID treatment facility, versus the standard (only) COVID19-treatment protocol, starting with 46 cases in each group, matched in terms of disease severity, of symptoms, and average cycle threshold value (CT-value) for the COVID PCR test on diagnosis.

Our findings showed statistically significant improvement in terms of a higher conversion rate to “COVID19-negative PCR” by five days of treatment (6/46 Vs 0/46), improvement in “symptoms severity” after two days of treatment, and less intubation and mortality (0/46 Vs 3/46) with all P-value < 0.05. There was also a trend of improvement in other outcome variables, though with no statistically significant difference; namely “shorter hospital stays,” “less progression in Oxygen requirements”, “less rate of plasma transfusion”, and better “gross extent of improvement”.

Our findings support a beneficial role in treating active cases (Disease) and anticipates better outcome should implemented earlier in course of the disease; thus, suggest a role in limiting the spread (Pandemic), as an additional preventive method. Additionally, we think the repeated reduction in the microbial load might have been sufficient to induce a strain in a possible viral-microbial interaction, resulting in slowing down of the disease progress.

## 3. Introduction / Background

The current treatment policies for COVID19 are rather “supportive”, including simultaneous approaches to maintain adequate oxygen supply, and preventing secondary infections, in addition to targeting the immunity itself, to optimize the outcome, and prevents overt reactions.

Considering the risk of exposure; the oral route is likely to pose and equal, if not a higher risk than the nasal; given the reported GIT symptoms in some cases, the lack of natural filtering capacity and the protection provided by the high levels of nitrous oxide produced (1, 2), which proved to inhibit viral replication, (3)

An existing state of dysbiosis reflects an impaired immune response, hence can facilitate developing the disease and subsequently the progression to worse outcomes.

It follow thus, the regular use of a potent mouthwash, that can consistently reduce the oral microbial load, including 2019-nCOV, aiding a faster recovery, as the immune systems is likely to struggle less to overcome the infection and consequently reduces the infection transmission rate. Additionally, this should lower the potential risk of other microbial superinfections. However, the sought benefits could also be achieved through another mechanism; invoking the evolutionary game theory, 2019-nCOV might be relying on an accomplice; be it a specific organism (e.g. bacterial species) or the state of dysbiosis in general, as the repeated and consistent reduction in the microbial load -oral microbiota and 2019-nCOV-might be sufficient to induce a strain in the viral-microbial interaction, inhibit reciprocal altruism, and negatively affecting the survival chance. The current literature indicates that 2019-nCOV is utilizing ACE2 receptors as access to the target cells (4), demonstrating the ability to induce both its upregulation through interferon-gene stimulation (5) possibly as a mean to increase its own replication, as well as down-regulation through direct binding (6) with effects that alter the natural lungs and gut microbiota in the direction of dysbiosis (7, 8), and an increased bacterial co-infection risk (9-16). ACE2 expression is identified in sites linked to the virus’s isolation and the disease complications, including the vascular endothelium, the respiratory system (10), and the intestines.

Additionally, both ACE and ACE2 have a structural similarity within their active-site region to other metalloproteases in some bacteria, including some of the URT pathogens; such as the M32 carboxypeptidase from the Bacillus subtilis (11), a probiotic and a possible agent of dental caries, and Paenibacillus sp. B38 with demonstrated ability to lower angiotensin II levels in mice (12), suggesting another possible direct viral -bacterial interaction. Thus, ACE2 can be utilized as a medium for microbial-viral interaction, and can be dialed up or down to maintain reciprocal altruism, especially among species with the potential to benefit from dysbiosis, as each side can affect the other, either directly or through inducing the host’s immune response.

### Mouth Wash and oral Microbiota

Chlorhexidine mouthwash (CHX) (0.12%-0.24%) is frequently used in dental practice, as it has a beneficial effect on controlling bacterial (13-17)(20-23) overgrowth, while the hydrogen peroxide (HPX) at a concentration of 3% demonstrated ability to reduce the viral load on surfaces by >4-log when applied for one-minute(17). HPX is produced naturally by both; the epithelial cells via superoxide dismutase enzyme, releasing ion superoxide, and by the oral bacteria itself, in effect to maintain the oral microecology(18, 19) through inducing oxidative stress similar to those triggered by viral infections; that stimulates a local innate response.(20) However, the combination with hydrogen peroxide (HPX) (1.5%-3%) proved to have a better outcome among users while maintaining the antibacterial efficiency as for CHX alone(37-39) within two weeks of use, even without specific dental hygiene instructions (40), as well as to control ventilator-associated pneumonia (VAP) measure (41, 42). Thus, this combination may have a great potential in controlling COVID19 (21), as it can induce an anti-viral response before the actual recognition of the viral antigens by the host immune cells.

## Objectives

### Primary Objectives

To determine the average recovery rate, in terms of nasopharyngeal swab test (COVID RT-PCR) for the intervention and control cases, after two weeks of treatment.

### Secondary Objectives

1. To determine the average Hospital stay for both; the study and control group
2. To determine the rate of COVID progression (deterioration) for the intervention and control groups.
3. To determine the average symptomatic improvement, using modified STAT-10 tool.
4. To determine 30-days Mortality rate amongst the intervention and control groups.

## Method

This is an investigator-initiated, randomized, unblinded, phase IV, clinical trial (ISRCTN**10197987: 05/10/2020**), has been approved and funded by IRB of the Hamad Medical Corporation’s Medical Research Center (MRC 05-106); the methods were carried out in accordance with relevant guidelines and regulations.

We consecutively recruited eligible patients of COVID-19, confirmed through combined Nasopharyngeal-Oropharyngeal swab PCR); who were admitted within 24 hours to a designated COVID-19 treatment facility in the State of Qatar (Hazm Mebaireek General Hospital), for either COVID-19 related complications, or those related to other comorbidities in COVID-19 positive cases. Medical records of eligible patients were reviewed, excluding those under 18 years of age, pregnant women, mental or cognitive impairment, maxillofacial injuries, those intubated or expected to be intubated within 24 hrs. Then eligible cases were counselled, and a written informed consent was obtained from all subjects, or their attending first degree relative for some of the geriatric cases; upon their request) recruited into the study.

As per the hospital’s protocol all cases have been reviewed by the independent COVID-team upon admission and, as demonstrated in Fig.1, assigned to “Clinical categories” (fig.1) based on symptomatology, clinical findings, and the results of blood and radiological tests. They are then started on treatment as per pre-specified protocols for the corresponding “categories” consistent with Communicable Diseases Center (CDC) Guidelines. These includes antivirals, antibiotics, steroids, in addition to hydroxychloroquine and convalescent plasma transfusion (where indicated).

Meanwhile for the “intervention” group, they received in addition to the standard protocol, three-times daily mouth rinse and gargles, for at least 30 seconds. This is comprised of 15 mls of mixed solution of 10 mls of 0.2% Chlorhexidine gluconate (oral rinse) plus 5 mls of 6% Hydrogen peroxide (to make up a final concentration of 2%). The solution constituents were mixed at bedside and presented to subjects. They were required to delays rinsing with tap water, eating or drinking to at least after 5 minutes of the mouthwash use. Since the underlying hypothesis was to consider the regular “repeated use” for long duration (2 weeks), those who had missed the intervention use for a day or more (>3 doses) were considered as a “drop-out”, and excluded from analysis.

Initially they were advised to use the mouthwash for one minute (not exceeding 2 minutes contact time with the oral cavity), however, due to difficulty of prolonged use given the high oxygen requirements, it was reduced to 30 seconds (for all cases). The first case was recruited on 10/09/2020, and the data collection completed by 30/10/2020.

The treatment was provided by the clinicians as per the hospital policy; while the research team was assigned only to prescribe the mouthwash, daily follow up through phone, remotely reviewing digital records, and swabs scheduling.

Study subjects of both groups had daily phone-based assessment; for evaluation of upper respiratory tract symptoms (using modified STAT-10 tool); in Supp.3.

From the digital medical chart, the following was updated daily

- The progress in clinical status (improving vs. deterioration).
  - Oxygen requirements: defined using two variables; the first is “on starting” treatment within first 24 hours of admission”, and the second is “progression in requirements” during the hospital stay.
  - Gross extent of improvement: derived to quantify the degree of improvement numerically, as “*the difference between the clinical category at the admission of the case, and its corresponding disposition*”. As both of the main components are 5-points categorical variables, with the “worse” is graded “higher”, the “gross extent of improvement” is calculated here by subtracting the former from the latter, in this sense for example, a case of “severe pneumonia” that gets to be discharged “home”, will have a higher score by two points, than a “mild” case that had the same disposition, with “negative” values indicate “improvement”, while “positive” values indicate progression/worsening of the disease.
- Treatment and Medications provided: the administration of “antibiotics, anti-virals and steroids”, sorted by the frequently used agents, as well as the pattern of intake (combination).
- Oropharyngeal and nasopharyngeal swab were collected on Day 5 and Day 15 of “starting treatment” in the intervention group and tested for COVID19 RT-PCR test.
- Disposition: Ranked categorically (best to worst) as: “Discharge home”, “Transfer to quarantine facility” ; “Extended hospital stay”, “Intubation”, and eventually “Death” (within 30 days of admission for discharged cases, or within the same hospitalization).

### Primary Outcome

Recovery rate (as per the latest update of CDC guidelines (Fig.1); based on “improvement in clinical symptoms, in addition to the results of COVID-PCR tested at 5 and 15 days of treatment.

### Secondary outcomes

- COVID progression (deterioration): defined as “need for intubation” or “death”.
- COVID improvement: defined as reallocation to lower level of care.
- Obtaining CT value > 30 in subsequent COVID RT-PCR test at days 5 and 15 of treatment.

## Results

From total admissions between 08/09/2020 – 01/10/2020, 147 COVID19 consecutive cases were screened, from which, a total 101 eligible patients were recruited and randomly allocated into an intervention (n =56) and control (n =46) groups as shown in the flow chart (Figure 2). At the end of the study, 43 and 44 cases were available for analyses in the intervention Vs. the control arm of the study respectively. Reasons for droputs from the study are given in supplementary material (SUPP 1), while the flow-chart for the study is in (SUPP 2). 92 cases were available for the first stage of analysis 46 in each group, which is marked by D5 swab; and 86 cases by D15 were available for the second swab; as the remaining cases could not undergo the tests as planned.

The characteristics of the study populations are given in table 1. There was no statistically significant difference between the two groups in terms of age (P = 0.89) and gender distributions, nor in terms of “average Cycle threshold (CT) value” on diagnosis PCR, the mean duration of symptoms prior to admission (fig.3), nor whether the onset of symptoms was before or after the diagnosis. About **19/92** (21%) have had no known comorbidities, with no statistically significant difference between the two groups in terms of morbidities as demonstrated in Table 1.

**Table 1:**
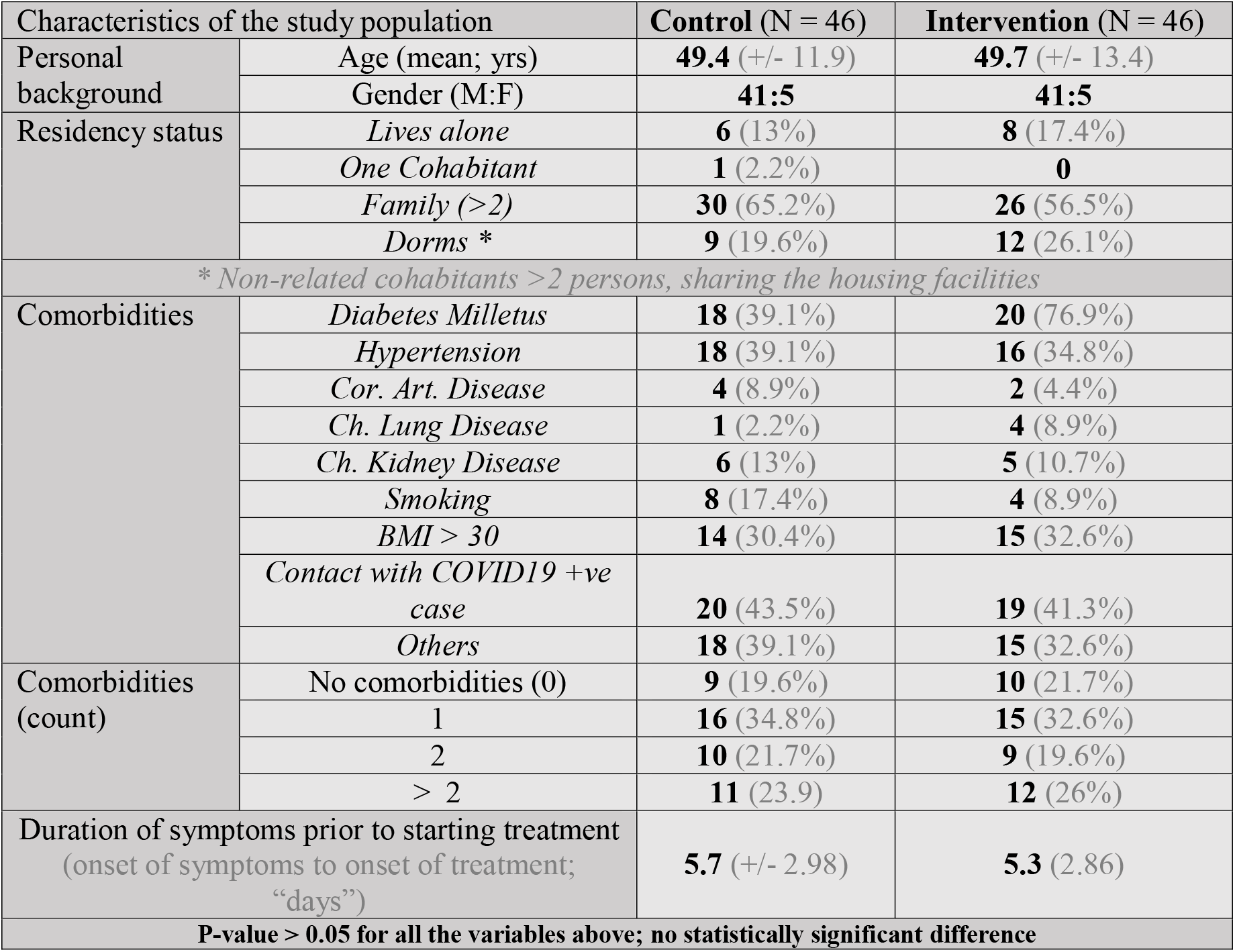
Characteristics of the sample population

### Clinical category on admission

Table 2 gives a summary of the clinical category / classification of patient cohorts within the two study arms. There was no statistically significant difference between the two groups.

**Table 2:**
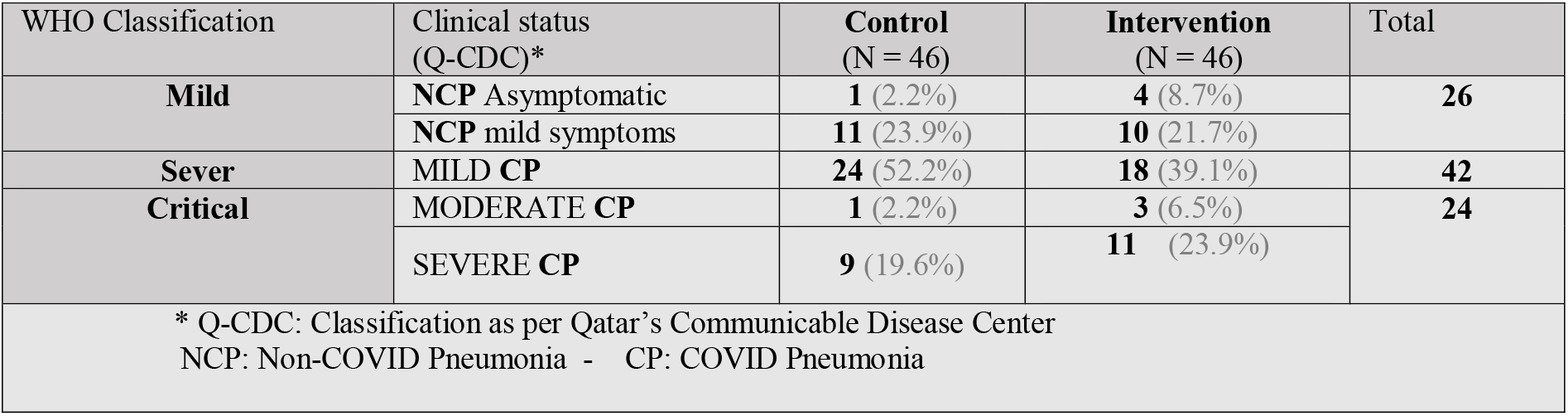
Clinical category on admission

### Treatment Received

There was no significant difference between the two groups in terms of treatment received (P-value > 0.05); with details in Table 3.

**Table 3:**
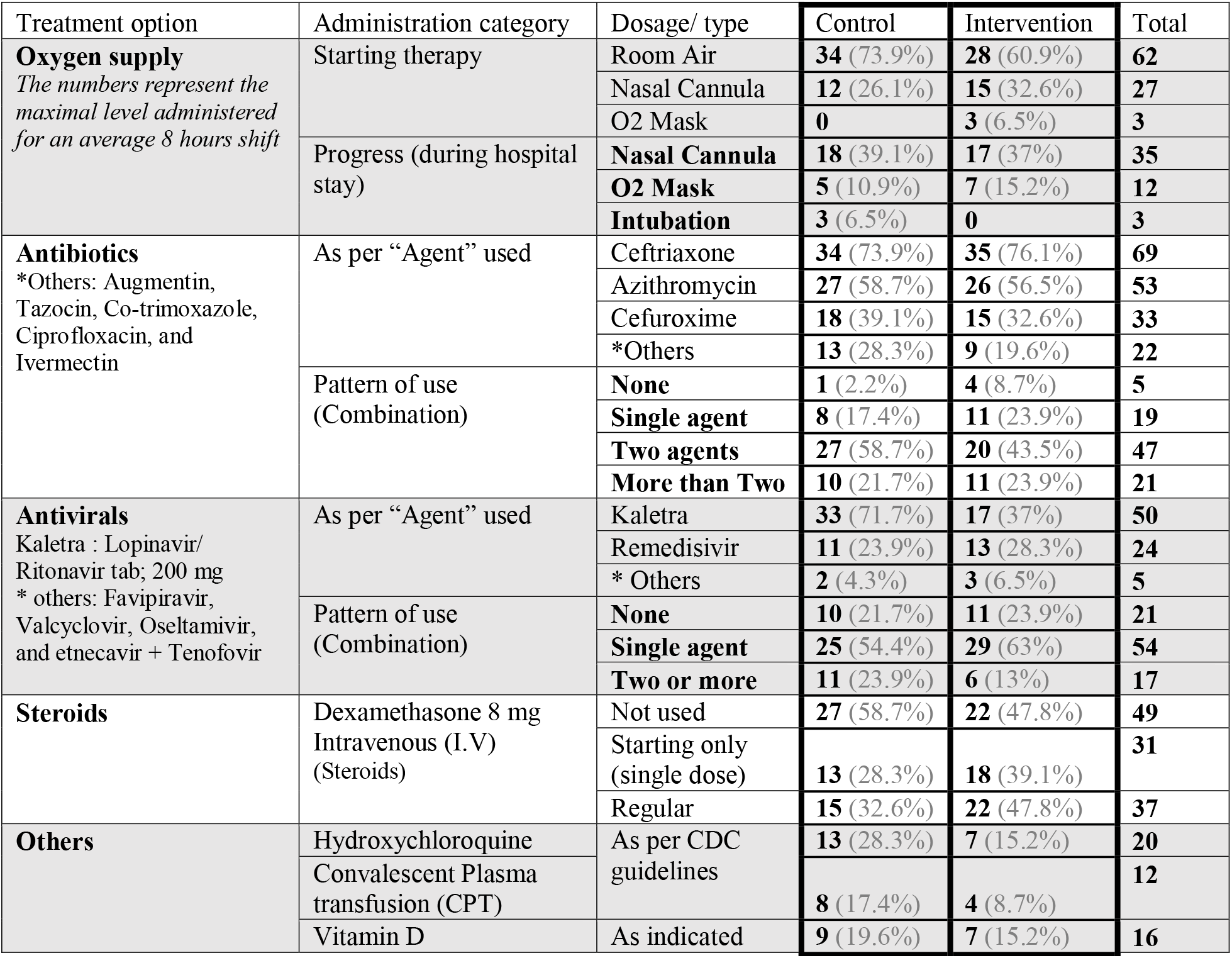
Treatment provided

### The use of the intervention (Mouthwash)

We found no statistically significant correlation between the “duration” of mouthwash use on one hand, and the COVID PCR outcome on Day 15, nor patient disposition, on the other hand.

### The COVID PCR swab test

As demonstrated in table 4; the test results are interpreted as three ordinal outcomes (“Negative”, “inconclusive” or “positive”).Those with “positive” test outcomes then expressed in terms of the average Cycle threshold (CT-value) of 5-units intervals, to demonstrate the variability within the category. Independent sample T-test done for the control and intervention groups per each corresponding “date”; the outcome discussed below.

**Table 4:**
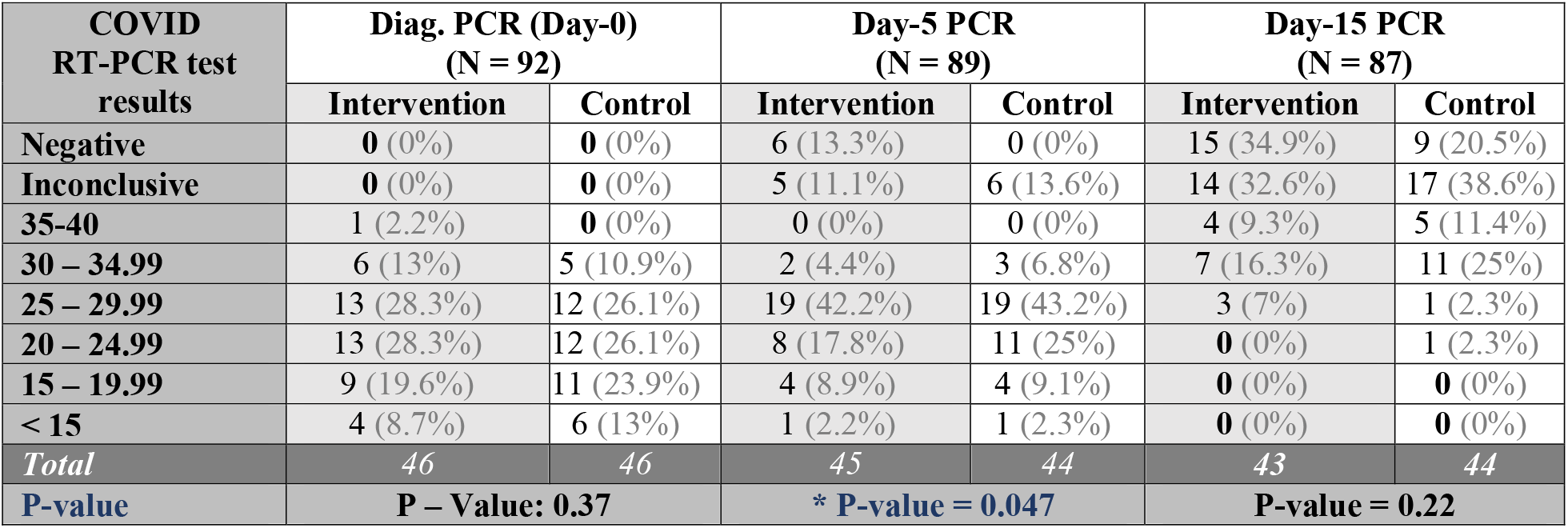
Results of COVID19-rtPCR for np/op Swabs; ad D:0 (diagnosis), Day-5 and Day-15 of treatment

### The COVID-19 syndrome outcome following Day-5 swab: (N = 89)

The 45 participants who received the drug intervention (*M* = 2.6, *SD* = 0.7) compared to the 44 participants in the control group (*M* = 2.9, *SD* = 0.4) demonstrated significantly better outcome *t*(87) = 4.1, P-value = .047 (fig.4). Of those who had negative results, 5/6 were symptomatic (1-6 days) prior to diagnosis (average 2.8 days), while the last one was only symptomatic 4 days after diagnosis (screening). Average CT-value (pre-intervention) for those whose swabs turned negative was 28.67 (95% CI: 21.77 – 35.58), Inconclusive 23.44 (95% CI: 20.65 – 26.22), while those that remained positive is 22.51 (95% CI: 21.09 – 23.92). Overall, there was a significant difference in terms of average CT-value on diagnosis between those with negative swabs on Day-5, and those who remained positive or had inconclusive results *t*(87) = 2.4, P-value = 0.017.

The average duration (days) of “symptoms” prior to starting treatment was variable with not no significant difference in the final estimates in those with negative (4.33 days), Inconclusive (7.27), and “positive” (5.42) swabs P-value: 0.083.

### COVID-19 syndrome outcomes byDay-15 swab (N = 87)

A total of 18 additional cases had “negative” PCR swabs by **day 15** (**9** on each study arm); this brings to a total to **15** cases amongst the 43 participants who received the drug intervention (*M* = 1.95, *SD* = 0.83) compared to 9 amongst the 44 participants in the control group (*M* = 2.2, *SD* = 0.8). (fig. 5) There was no statistically significant difference between the two groups (*t*(85) = **1.5**, P-value = **0.22)**. There was no statistically significant difference between the two groups in terms of PCR results on Day 15 (for those remained positive), and duration of symptoms prior to the diagnosis, the average duration of symptoms prior to starting treatment, nor the hospital stay duration; P-value: > 0.05.

### Hospital Stay

As demonstrated in Figure 6 (FiguresTables); the mean duration of hospital stays was comparable between the two groups (intervention mean **8.11** (95% CI: 6.19 -10.02), vs. Control: mean **9.43** (95% CI: 7.15 -11.72); with the extended hospital stay greatly affected by existing comorbidities as well as other COVID-related complications.

### Final patient disposition (N = 92)

As demonstrated in Figure 7 (FiguresTables); cases allocated to the intervention group had higher discharge and transfer rates, and less extended hospital stays, need for intubation, and overall mortality compared to their control cohorts (P-value = **0.04981)**.

Three cases (**3/46; 6.5%**) were intubated in the “control group”, of them two passed away, yielding the mortality within this group (**2/46; 4.4%**) during the hospitalization; one at day **18**, and the other on day **35** of the hospital stay; the latter case was discharged, then admitted again (hence the total duration > 30 days) to another facility two days later due to exacerbation of symptoms, intubated again, diagnosed as multiorgan failure within 3 days, then passed away after 6 days. A third case from the control group has passed away at the 54th day of hospital stay; by then has already been intubated for six weeks. No mortalities reported among the intervention group for at least 4 weeks from discharge, including those dropped-out.

### Gross extent of improvement

(table 5; supp.) Being a derivative of two categories, the intervention group has a better score as shown in figure 8 (FiguresTables), but not statistically significant (P-value > 0.05).

### Symptomatic improvement

Using modified STAT-10 score for sore throat; less score represents “less symptoms severity”, and on daily bases, used to indicate improvement. There was a significant difference between the two groups in the “average daily score” starting from Day 3, Day 4 and Day 5; as shown in figure 9 (FiguresTables).

## Discussion

Despite of the small sample size, when both “intervention” and “control” groups are matched in terms of age, comorbidities, duration of symptoms prior to hospitalization, clinical category on admission, the average CT-value of the COVID-PCR on diagnosis, and different components of the treatment protocol used; the regular use of mouthwash in cases hospitalized for COVID19 seems to improve the outcome; this finding extends beyond the therapeutic value in a symptomatic case, as it implies less probability of infection transmission, as evident by the statistically significant earlier conversion to “COVID-negative” by 5 days of treatment, improvement in symptoms after 2 days of treatment, and the “better” disposition; with no intubation nor mortality.

There was also a trend of improvement in other outcome variables, though with no significant statistical difference; namely “hospital stay”, “less progression in Oxygen requirements”, “less rate of plasma transfusion”, and the “gross extent of improvement” in terms of disposition and the clinical category on admission.

The observed improvement suggests an additive value to the treatment protocols for the hospitalized COVID19 cases, which takes the credit for the overall improvement in both groups. The frequency of the intervention “mouthwash” use as counted during the hospital stay period, was a quantitative rather than a qualitative assessment; it was linked primarily to the hospital stay duration, and frequently reported, especially among the elderly and those with higher oxygen requirements, to have poor compliance with either the method, i.e. the “instructions to gargle and move the solution within the cavity”, or the “duration of the use up to **30** seconds”; we do consider this as a limitation to be addressed in future studies or implementation, although there’s no consensus in the literature to support the significance of the “quality of use”, duration” nor “frequency” as requirements for certain outcome.

Our findings in terms of intervention use within the intervention group, are clearly against a possible dose-related negative-conversion, at least beyond the 5^th^ day of treatment, this supported by the absence of a linear relationship between the frequency of intervention use and the average CT-value and, at least in the term of duration, yet in the view of improvement in other parameters, such absence in linear relationship favors possible interference in a necessary the viral-microbial interaction for the disease progression, which in lab settings, is likely to display as a disproportional reduction in the viral cytopathological activity (through culture), to the sequentially decreasing viral load (by Ct-value).

Taking this into account, we advise to use the intervention earlier in the disease course, in order to get optimal outcome, including the pre-symptomatic and asymptomatic cases, and hence by extension, to those “exposed”. Since the exposure is usually discerned in retrospect, it seems logical to implement the mouthwash use in the community, as an additional “preventive” measure.

We advise repeating this study in the view of different treatment protocols implemented worldwide; given the variation in treatment options. However, in the view of the ongoing pandemic, we suggest endorsing such low-cost intervention -regular use of portent mouthwash-for both, hospitalized COVID19 cases, as well the public, especially in closed communities.

## Conclusion

Within the current constituents’ concentration and frequency of use, the regular use of potent mouthwash solutions seems to accelerate the recovery of COVID19 and seems to have no linear relationship with the duration of use. This observed improvement suggests better potential in an earlier stage of the disease, as an addition to the treatment protocols for the hospitalized COVID19 cases, especially for high-risk populations. By extension, as the solution constituents are available commercially as over the counter items, we recommend its use as an additional pandemic control measure, especially for closed communities, such as nursing home and similar long-term facilities, schools, sports training facilities, as well as prisons and army dormitories.

Finally, we strongly advise repeating this study in larger sample and with different treatment protocols, to quantify the extent of improvement. However, this shouldn’t be a reason to delay endorsing its use in the community.

## Supporting information

Methods (detailed)

## Data Availability

The Data sheet referenced in the manuscript is to be shared only with a publishing journal, after signing a Data sharing agreement

https://www.isrctn.com/ISRCTN10197987

## 2. Abbreviations and Acronyms

RTI: Respiratory Tract Infection
COVID19: novel Coronavirus disease 2019
2019-nCOV: the novel coronavirus that causes severe acute respiratory illness/ Syndrome
RT-PCR: Reverse transcription Polymerase Chain reaction
CT: Cycle threshold value; the number of amplification cycles required to reach a fixed (fluorescent) signal to cross the threshold (i.e., exceeds background level).

## Additional information

### Authors contribution

khalid Mukhtar ^1^ (First & corresponding Author):

Formulating the hypothesis, Methodology, proposal, methodology and manuscript writing, funding acquisition, data collection supervision, data analysis, results interpretation.

### Data Curation

Mohammed F. Khudair ^7^, Housamaddeen Al Farhan ^6^, Ahmed Hassib ^11^, Rumaisa Mohamed Elmustafa ^12^, Wesal Elhadary ^13^, and Morwan Abdulkarim ^14^

### Supervision (data curation)

Suelen Qassim ^2^, Shaikha Ali Al Qahtani ^3^, Mohamed Mohamedali ^5^, Rajvir Singh ^15^

### Writing (original draft, review & editing)

Khalid Mukhtar^1^ and Mohamed Ibn-Masud Danjuma ^4^

### Project administration

Abdel Rehim El Tayeh ^8^, Mohammed Al-Dosari ^9^, Mohamed Elhassan Babiker ^10^, Muna Al.Maslamani ^16^

### Transparency Statement

The lead authors affirms that the manuscript is an honest, accurate, and transparent account of the study being reported; that no important aspects of the study -relevant to this publication-have been omitted; and that any discrepancies from the study as planned (and, if relevant, registered) have been explained

### Data availability Statement

The data that support the findings of this study are uploaded upon submission, and will be made available for reviewer through a link to an open repository upon study approval.

