## Supplementary material for "A randomized trial on the regular use of potent mouthwash in COVID-19 treatment": Methods (detailed)

### Method

This is an investigator-initiated, randomized, unblinded, phase IV, clinical trial (ISRCTN**10197987: 05/10/2020**), has been approved and funded by IRB of the Hamad Medical Corporation’s Medical Research Center (MRC 05-106); the methods were carried out in accordance with relevant guidelines and regulations. Neither the patients nor the public WERE NOT involved in the design, or conduct, or reporting, or dissemination plans of our research.

We consecutively recruited eligible patients (with COVID-19 confirmed through combined Nasopharyngeal-Oropharyngeal swab PCR) ;who were admitted within 24 hours to the largest COVID-19 treatment facility in the State of Qatar (Hazm Mebaireek General Hospital) , for either COVID-19 related complications, or those related to other comorbidities in addition to COVID-19 positive cases. Medical records of eligible patients were reviewed, excluding those under 18 years of age, pregnant women, mental or cognitive impairment, maxillofacial injuries, those intubated or expected to be intubated within 24 hrs.

The eligible cases were then counselled, and a written informed consent was obtained from all subjects, or their attending first degree relative for some of the geriatric cases; upon their request) recruited into the study.

As per the hospital’s protocol/usual standard of care, all cases have been reviewed by the COVID-team physicians upon admission and assigned to “Clinical categories**” based on symptomatology, clinical findings, and the results of blood and radiological tests. They are then commenced on treatment cocktails as per pre-specified protocols for the corresponding “categories” consistent with Communicable Diseases Center (CDC) Guidelines. These includes antivirals, antibiotics, steroids, in addition to hydroxychloroquine and convalescent plasma transfusion (where indicated).

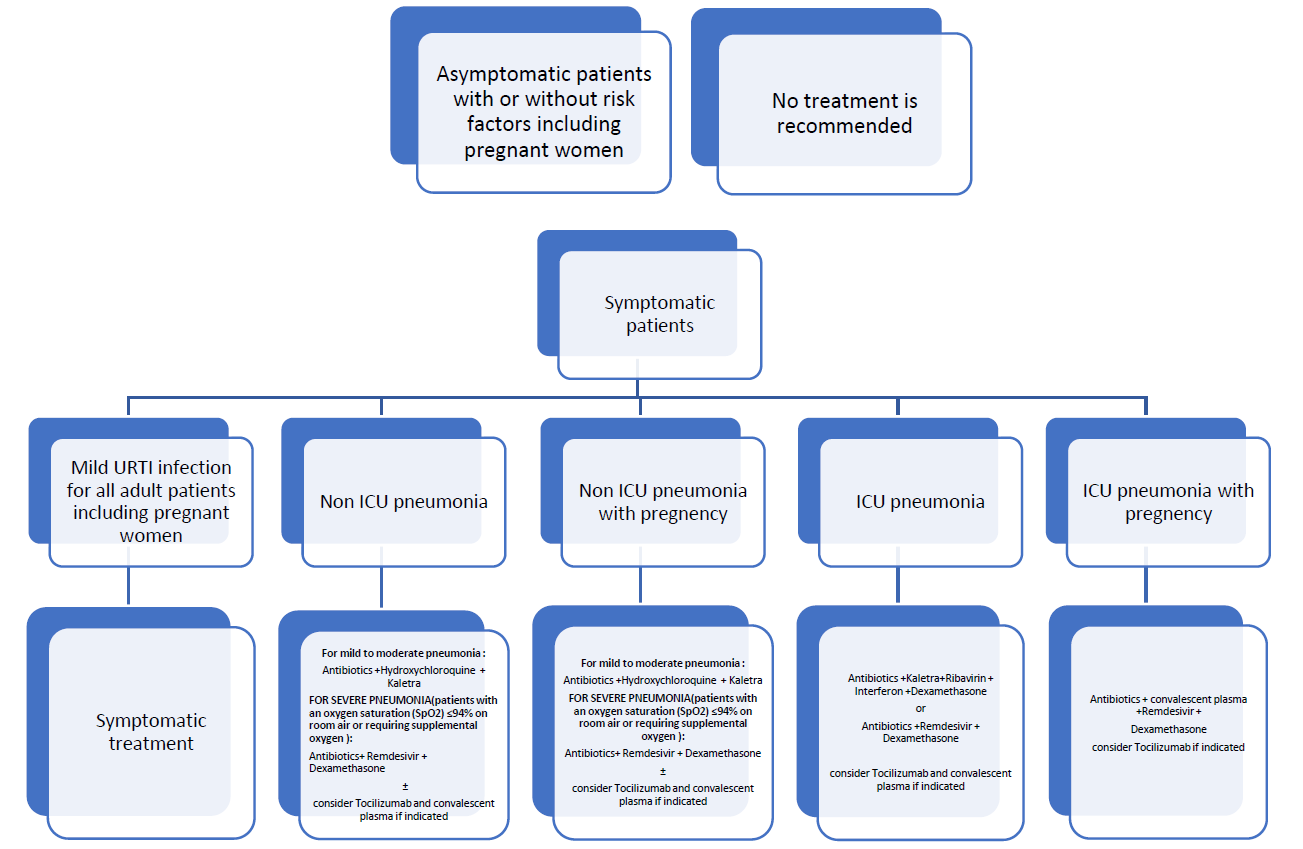

Figure 1: ** (Prepared by HMC CDC COVID-19 Scientific Committee (Doha-Qatar), version 10 (06/09/2020))

For the study , once potential eligible were cases identified, they were first contacted through phone calls, and had the study rationale and methodology explained to them (as per the “pre-consent phone-call script”). Once they agreed to participate, they were be required to sign a consent form. Given the pattern of admissions, candidates were recruited in daily quotas, that were assigned through block randomization to two groups; an intervention and control arm. To compensate for the inevitable “discontinued use” and “drop out”, more slots were allocated to the “intervention” group as more patients accrue.

Both groups were receiving standard COVID treatment as per the CDC protocol, while for the “intervention” group, additionally three-times daily mouth rinse and gargles, for at least 30 seconds. This is comprised of 15 mls of mixed solution of 10 mls of 0.2% Chlorhexidine gluconate (oral rinse) plus 5 mls of 6% Hydrogen peroxide (to make up a final concentration of 2%). The solution constituents were mixed at bedside and presented to subjects. They are required to delays rinsing with tap water, eating or drinking to at least after 5 minutes of the mouthwash use. Since the underlying hypothesis was to consider regular “repeated use” for long duration (2 weeks), those who had missed the intervention use for a day or more (>3 doses) were considered as a “drop-out”, and excluded from analysis.

The treatment period ranged between one to 15 days; as it was provided three times a day, and as it was calculated for the “in-patient” period only, as documented by the assigned nurse.

Initially they were advised to use the mouthwash for one minute (not exceeding 2 minutes contact time with the oral cavity), however, due to difficulty of prolonged use given the high oxygen requirements, it was reduced to 30 seconds (for all cases).

The first case was recruited on 10/09/2020, and the data collection completed by 30/10/2020; however, those who were still in-patient were reviewed again in 4 weeks to confirm the final outcome, although they were indicated as “extended hospital stay” in the results of this paper.

**Follow up**

The treatment was provided by the clinicians as per the hospital policy; while the research team was assigned only to prescribe the mouthwash, daily follow up through phone, and schedule the swabs.

Study subjects both groups had daily phone-based assessment; for evaluation of upper respiratory tract symptoms of using modified STAT-10 tool.

From the digital medical chart, the following was updated daily

- Clinical status including vital signs, oxygen supply requirements, progress in clinical status (improving vs. deterioration).
  - Oxygen requirements were defined using two variables; the first is “on starting” treatment within first 24 hours of admission”, and again for “progression in requirements” during the hospital stay.
  - Gross extent of improvement: We derived this variable to quantify the degree of improvement numerically, defined as “*the difference between the clinical category at the admission of the case, and the corresponding disposition*”. As both of the main components are 5-points categorical variables, with the “worse” is graded “higher”, the “gross extent of improvement” is calculated here by subtracting the former from the latter, in this sense for example, a case of “severe pneumonia” that gets to be discharged “home”, will have a higher score by two points, than a “mild” case that had the same disposition, with “negative” values indicate “improvement”, while “positive” values indicate progression/worsening of the disease.
- Treatment and Medications provided: Were reported from Medications charts for the “in-patient” only. For the “antibiotics, anti-virals and steroids”, they were presented in two distinct ways; sorted by the frequently used agents, as well as the pattern of intake (combination).
- Oropharyngeal and nasopharyngeal swab were collected on Day 5 and Day 15 of “starting treatment” in the intervention group and tested for COVID19 RT-PCR test.
- Disposition: list of actions taken according to the change in the clinical status spectrum, used as a proxy for the “Case improvement / progression”, in relation to the “treatment protocol”. Ranked categorically as:
  - Discharge home: which represents the best outcome; additional medications may be prescribed as indicated, and instructed for 5 days of self-isolation
  - Transfer to quarantine; the second-best option, as such facilities are being prepared by facility doctors and nursing staff, this is the preferred decision when medical supervision is required in addition to “isolation”.
  - Extended hospital stay: It represents either continuum of the same hospital stay, transfer to another hospital, or readmission within 48 hours; as defined per the purpose of the study. It’s rather neutral in the improvement-progression spectrum.
  - Intubation: Being a unique identification for higher oxygen requirements, used as a proxy for “disease progression / deterioration”; as the entire inpatients’ section of the study area (hospital) is upgraded to High-dependency unit, with no clear distinctions between “ICU” and “ward” in terms of physical boundaries nor most of the treatment guidelines.
  - Death: represented the worst outcome in the disposition scale and considered only if reported within 30 days of admission.

**Primary Outcome**

- Recovery rate (as per the latest update of CDC guidelines **); based on “improvement in clinical symptoms, in addition to the results of COVID-PCR tested at 5 and 15 days of treatment.

**Secondary outcomes**

- COVID progression: defined as “need for intubation” or “death”. The onset is taken as “date where the deterioration was first reported.
- COVID improvement: defined as reallocation to lower level of care and or discharge to specialized facility or unit. The onset is taken as “date where the improvement was first reported, and consequently followed by an actual transfer / care downgrade within 3 days”.
- Obtaining CT value > 30 in subsequent COVID RT-PCR test at days 5 and 15 of treatment.

**Swabs details**

The brand and specification of “swabs” used for Qatar samples.

1.    UTNFS - VIRAL COLLECTION, PRESERVATION AND TRANSPORT MEDIUM KIT . ( NASOPHARYNGEAL SWAB + OROPHARYNGEL SEAB) +2 ML UTM TUBE. Mfr : noble bio korea, catalogue : UTNFS-3B-2

2.    UTM Plus 3ML NP Flocked Swb + Regular Flocked Swab. Mfr: huachenyang technologies, catalogue : CY-F002-21

3.    UTM Plus 3ML NP Flocked Swb + Regular Flocked Swab. Mfr: Copan Italia, catalogue : 321C.

**Swabs processing:**

Combined nasopharyngeal and oropharyngeal swabs (Copan Diagnostics Inc, Italy) when collected are placed in Universal Transport Medium (UTM). Aliquots of UTM are either extracted on the  QIAsymphony platform (Qiagen, USA) and tested by RT-PCR with the Thermo Fisher TaqPath COVID-19 RT-PCR Kit (Thermo Fisher, USA), targeting the S, N  and ORF-1a/b E-genes, or loaded directly on to a Roche Cobas® 6800 and assayed with the Cobas® SARS-CoV-2 Test (Roche, Switzerland) targeting the ORF-1a/b and E-gene regions of SARS-CoV-2.

This study has been approved by IRB of the Hamad Medical Corporation’s Medical Research Center (MRC 05-106); the trial registration number: **ISRCTN10197987**

Transparency Statement:

The lead authors affirms that the manuscript is an honest, accurate, and transparent account of the study being reported; that no important aspects of the study -relevant to this publication- have been omitted; and that any discrepancies from the study as planned (and, if relevant, registered) have been explained

The formal written dissemination of the study results to these groups is not possible/applicable; however, we verbally communicated the main findings to some of the cases who showed interest.
