## Supplementary material for "A randomized trial on the regular use of potent mouthwash in COVID-19 treatment": Figures

| Table of contents | | |
| --- | --- | --- |
| Page | Item No. | Title |
| **Figures & Graphs** | | |
| 2 | Figure 1 | Clinical triaging and management for COVID19 cases; (Prepared by HMC CDC COVID-19 Scientific Committee (Doha-Qatar), version 10 (06/09/2020)) |
| 3 | Figure 2 | Study flow chart |
| 4 | Figure 3 | The period (in days; Y-axis) between onset of symptoms to starting treatment. The X-axis represents the frequency/cases count. (N = 92) |
| 4 | Figure 4 | COVID19 RT-PCR outcome at Day 5 |
| 5 | Figure 5 | COVID-19 RT-PCR outcome at Day 15 |
| 5 | Figure 6 | Hospital stay duration (weeks) |
| 6 | Figure 7 | Final patient disposition at the end of the study |
| 6 | Figure 8 | Gross extent of improvement "Disposition" in relation to the "clinical category on admission" |
| 7 | Figure 9 | Symptomatic improvement in the first 5 days; assessed using modified STAT-10 tool. |

Table 1: Characteristics of the

Figures:

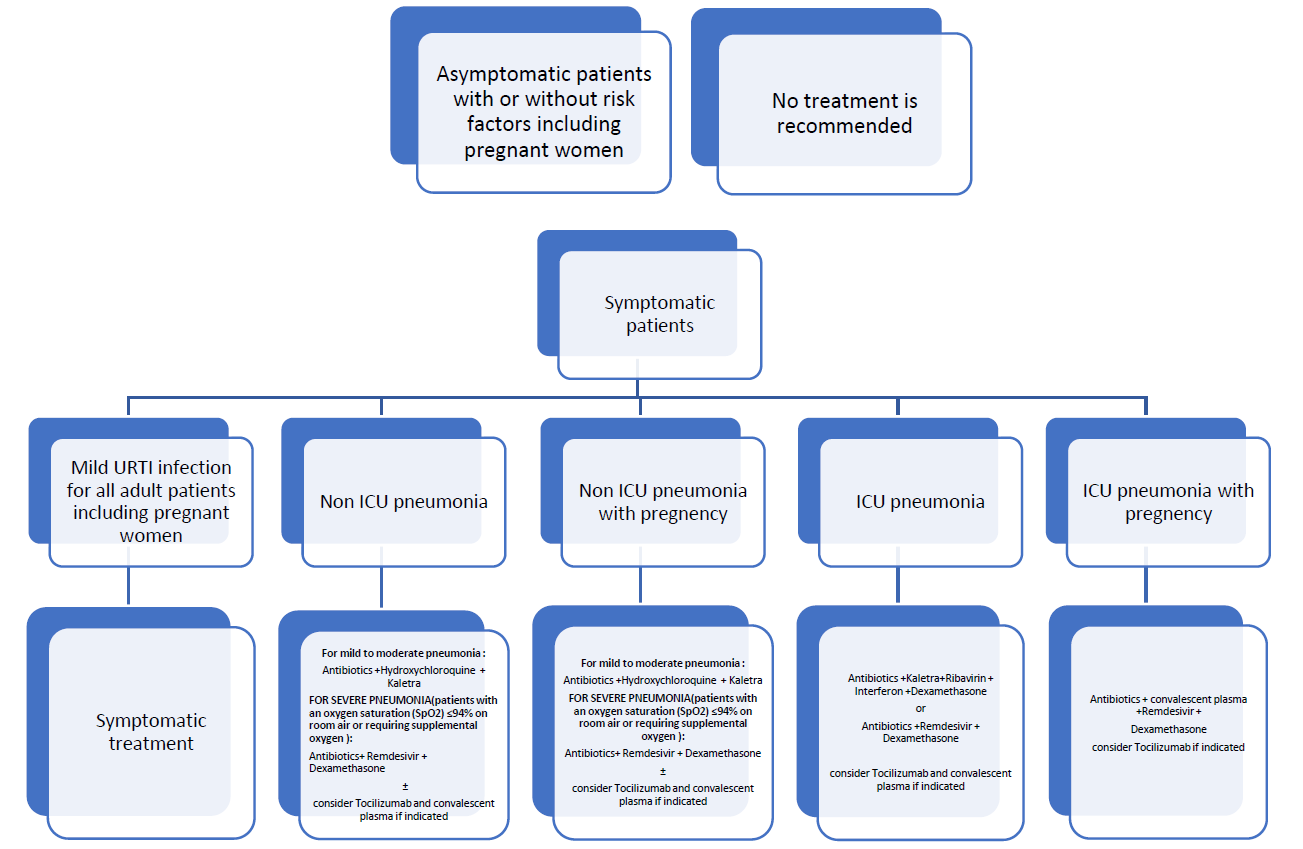

Figure 1: ** (Prepared by HMC CDC COVID-19 Scientific Committee (Doha-Qatar), version 10 (06/09/2020))

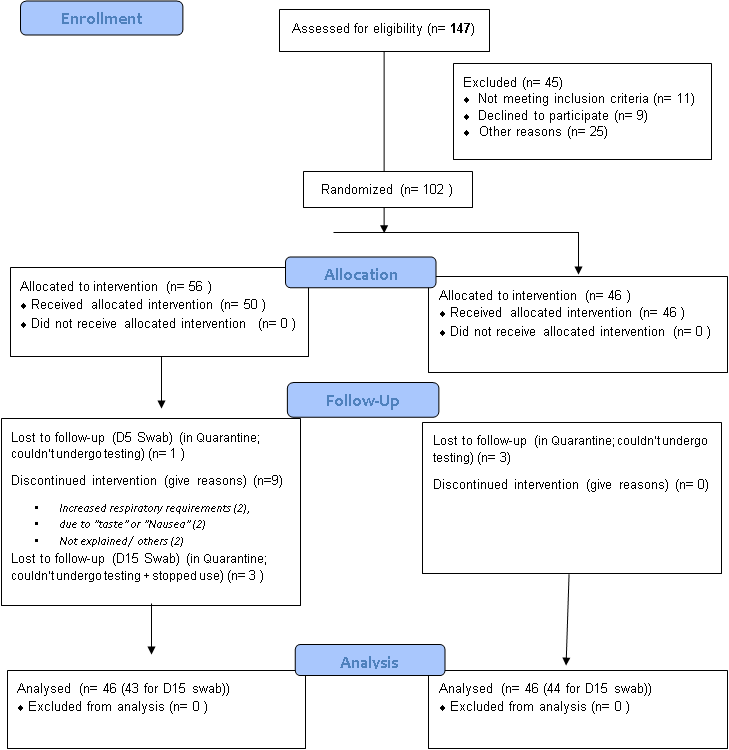

Figure 2: Study flow Chart

|  | Control | Intervention |
| --- | --- | --- |
| 1 | 0 | 4 |
| 2 | 7 | 2 |
| 2.68313 | 5 | 6 |
| 4 | 9 | 8 |
| 5 | 5 | 6 |
| 5.58852 | 5 | 6 |
| 7 | 4 | 6 |
| 8 | 2 | 3 |
| 9 | 5 | 1 |
| 10 | 2 | 0 |
| 10.669 | 1 | 3 |
| 12 | 1 | 0 |
| 14 | 1 | 1 |

Figure 3: The period (in days; Y-axis) between onset of symptoms to starting treatment. The X-axis represents the frequency/cases count. (N = 92)

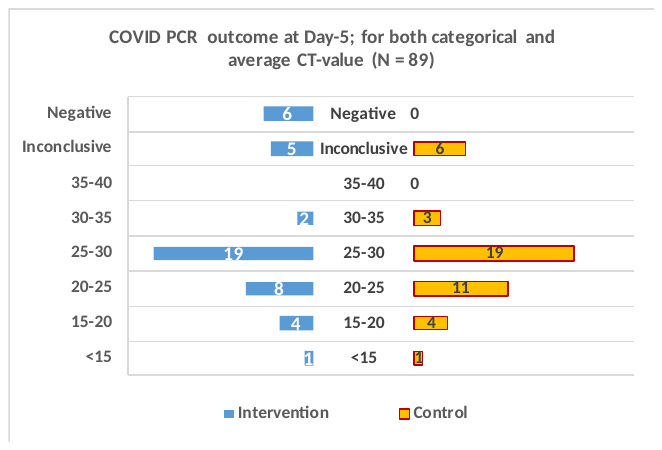

Figure 4: COVID19 RT-PCR outcome at Day 5

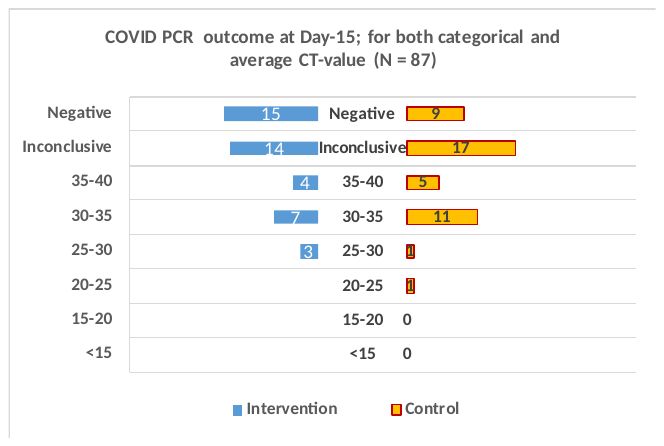

Figure 5: COVID-19 RT-PCR outcome at Day 15

N = 93

|  | Pinter | Intervention | Gap | Control | Pcontr |
| --- | --- | --- | --- | --- | --- |
| <1 Week | 3 | 27 | 12 | 26 | 4 |
| 1-2 Weeks | 16 | 14 | 12 | 11 | 19 |
| 2-4 Weeks | 25 | 5 | 12 | 6 | 24 |
| > 4 Weeks | 29 | 1 | 12 | 3 | 27 |

Figure 6: Hospital stay duration (weeks); 75% of the intervention group discharged by the 9^th^ day, in contrast to the 11^th^ day for the same percentage in the control group.

Cases disposition (N = 92)

|  | Pint | Intervention | Gap | Control | Pcont |
| --- | --- | --- | --- | --- | --- |
| Discharged (Home) | 2 | 28 | 14 | 22 | 3 |
| Quaratine facility | 14 | 16 | 14 | 17 | 8 |
| Inpatient | 28 | 2 | 14 | 4 | 21 |
| Intubated | 30 | 0 | 14 | 3 | 22 |
| Death | 30 | 0 | 14 | 2 | 23 |

Figure 7: Final patient disposition at the end of the study

Improvement Extent (N = 92) Disposition - Clinical stage

|  | Cotrol | Intervention |
| --- | --- | --- |
| -4 | 3 | 5 |
| -3 | 4 | 6 |
| -2 | 13 | 16 |
| -1 | 16 | 10 |
| 0 | 7 | 8 |
| 1 | 3 | 1 |

Figure 8: Gross extent of improvement "Disposition" in relation to the "clinical category on admission"

Symptomatic Improvement)

|  | Pint | Intervention | Gap | Control | Pcont |
| --- | --- | --- | --- | --- | --- |
| Day 1 | 6.4 | 13.6 | 8 | 16.3 | 3.7 |
| Day 2 | 9 | 11 | 8 | 13.8 | 1.2 |
| Day 3 | 12.4 | 7.6 | 8 | 14.8 | 0.2 |
| Day 4 | 14.4 | 5.6 | 8 | 11.2 | 3.8 |
| Day 5 | 16.1 | 3.9 | 8 | 7.6 | 12.4 |

Figure 9: Symptomatic improvement in the first 5 days; assessed using modified STAT-10 tool.
