## Supplementary material for "A randomized trial on the regular use of potent mouthwash in COVID-19 treatment": Tables

| Table of contents | | |
| --- | --- | --- |
| Page | Item No. | Title |
| **Tables** | | |
| 2 | Table 1 | Characteristics of the study population |
| 2 | Table 2 | Clinical category on admission |
| 3 | Table 3 | Treatment provided |
| 4 | Table 4 | Results of COVID19-rtPCR for np/op Swabs; ad D:0 (diagnosis), Day-5 and Day-15 of treatment |

Table 1: Characteristics of the study population

| Characteristics of the study population | | **Control** (N = 46) | **Intervention** (N = 46) |
| --- | --- | --- | --- |
| Personal background | Age (mean; yrs) | **49.4** (+/- 11.9) | **49.7** (+/- 13.4) |
|  | Gender (M:F) | **41:5** | **41:5** |
| Residency status | Lives alone | **6** (13%) | **8** (17.4%) |
|  | One Cohabitant | **1** (2.2%) | **0** |
|  | Family (>2) | **30** (65.2%) | **26** (56.5%) |
|  | Dorms * | **9** (19.6%) | **12** (26.1%) |
| ** Non-related cohabitants >2 persons, sharing the housing facilities* | | | |
| Comorbidities | Diabetes Mellitus | **18** (39.1%) | **20** (76.9%) |
|  | Hypertension | **18** (39.1%) | **16** (34.8%) |
|  | Cor. Art. Disease | **4** (8.9%) | **2** (4.4%) |
|  | Ch. Lung Disease | **1** (2.2%) | **4** (8.9%) |
|  | Ch. Kidney Disease | **6** (13%) | **5** (10.7%) |
|  | Smoking | **8** (17.4%) | **4** (8.9%) |
|  | BMI > 30 | **14** (30.4%) | **15** (32.6%) |
|  | Contact with COVID19 +ve case | **20** (43.5%) | **19** (41.3%) |
|  | Others | **18** (39.1%) | **15** (32.6%) |
| Comorbidities (count) | No comorbidities (0) | **9** (19.6%) | **10** (21.7%) |
|  | 1 | **16** (34.8%) | **15** (32.6%) |
|  | 2 | **10** (21.7%) | **9** (19.6%) |
|  | > 2 | **11** (23.9) | **12** (26%) |
| Duration of symptoms prior to starting treatment (onset of symptoms to onset of treatment; “days”) | | **5.7** (+/- 2.98) | **5.3** (2.86) |
| **P-value > 0.05 for all the variables above; no statistically significant difference** | | | |

Table 2: Clinical category on admission

| WHO Classification | Clinical status  (Q-CDC)* | **Control** (  N = 46) | | **Intervention**  (N = 46) | Total |
| --- | --- | --- | --- | --- | --- |
| Mild | **NCP** Asymptomatic | **1** (2.2%) | **4** (8.7%) | | **26** |
|  | **NCP** mild symptoms | **11** (23.9%) | **10** (21.7%) | |  |
| Sever | MILD **CP** | **24** (52.2%) | **18** (39.1%) | | **42** |
| Critical | MODERATE **CP** | **1** (2.2%) | **3** (6.5%) | | **24** |
|  | SEVERE **CP** | **9** (19.6%) | 11 (23.9%) | |  |
| * Q-CDC: Classification as per Qatar Communicable Disease Center   - NCP: Non-COVID Pneumonia, CP: COVID Pneumonia | | | | | |

Table 3: Treatment provided

| Treatment option | Administration category | Dosage/ type | Control | Intervention | Total |
| --- | --- | --- | --- | --- | --- |
| **Oxygen supply**  *The figures represent the maximal level administered for an average 8 hours shift* | Starting therapy | Room Air | **34** (73.9%) | **28** (60.9%) | **62** |
|  |  | Nasal Cannula | **12** (26.1%) | **15** (32.6%) | **27** |
|  |  | O2 Mask | **0** | **3** (6.5%) | **3** |
|  | Progress (during hospital stay) | **Nasal Cannula** | **18** (39.1%) | **17** (37%) | **35** |
|  |  | **O2 Mask** | **5** (10.9%) | **7** (15.2%) | **12** |
|  |  | **Intubation** | **3** (6.5%) | **0** | **3** |
| **Antibiotics**  *Others: Augmentin, Tazocin, Co-trimoxazole, Ciprofloxacin, and Ivermectin | As per “Agent” used | Ceftriaxone | **34** (73.9%) | **35** (76.1%) | **69** |
|  |  | Azithromycin | **27** (58.7%) | **26** (56.5%) | **53** |
|  |  | Cefuroxime | **18** (39.1%) | **15** (32.6%) | **33** |
|  |  | *Others | **13** (28.3%) | **9** (19.6%) | **22** |
|  | Pattern of use (Combination) | **None** | **1** (2.2%) | **4** (8.7%) | **5** |
|  |  | **Single agent** | **8** (17.4%) | **11** (23.9%) | **19** |
|  |  | **Two agents** | **27** (58.7%) | **20** (43.5%) | **47** |
|  |  | **More than Two** | **10** (21.7%) | **11** (23.9%) | **21** |
| Antivirals  Kaletra : Lopinavir/ Ritonavir tab; 200 mg  * others: Favipiravir, Valcyclovir, Oseltamivir, and etnecavir + Tenofovir | As per “Agent” used | Kaletra | **33** (71.7%) | **17** (37%) | **50** |
|  |  | Remedisivir | **11** (23.9%) | **13** (28.3%) | **24** |
|  |  | * Others | **2** (4.3%) | **3** (6.5%) | **5** |
|  | Pattern of use (Combination) | **None** | **10** (21.7%) | **11** (23.9%) | **21** |
|  |  | **Single agent** | **25** (54.4%) | **29** (63%) | **54** |
|  |  | **Two or more** | **11** (23.9%) | **6** (13%) | **17** |
| Steroids | Dexamethasone 8 mg Intravenous (I.V)  (Steroids) | Not used | **27** (58.7%) | **22** (47.8%) | **49** |
|  |  | Starting only (single dose) | **13** (28.3%) | **18** (39.1%) | **31** |
|  |  | Regular | **15** (32.6%) | **22** (47.8%) | **37** |
| Others | Hydroxychloroquine | As per CDC guidelines | **13** (28.3%) | **7** (15.2%) | **20** |
|  | Convalescent Plasma transfusion (CPT) |  | **8** (17.4%) | **4** (8.7%) | **12** |
|  | Vitamin D | As indicated | **9** (19.6%) | **7** (15.2%) | **16** |

Table 4: Results of COVID19-rtPCR for np/op Swabs; ad D:0 (diagnosis), Day-5 and Day-15 of treatment:

| **COVID**  **RT-PCR test results** | **Diag. PCR (Day-0)**  **(N = 92)** | | **Day-5 PCR**  **(N = 89)** | | **Day-15 PCR**  **(N = 87)** | |
| --- | --- | --- | --- | --- | --- | --- |
|  | **Intervention** | **Control** | **Intervention** | **Control** | **Intervention** | **Control** |
| **Negative** | **0** (0%) | **0** (0%) | 6 (13.3%) | 0 (0%) | 15 (34.9%) | 9 (20.5%) |
| **Inconclusive** | **0** (0%) | **0** (0%) | 5 (11.1%) | 6 (13.6%) | 14 (32.6%) | 17 (38.6%) |
| **35-40** | 1 (2.2%) | **0** (0%) | 0 (0%) | 0 (0%) | 4 (9.3%) | 5 (11.4%) |
| **30 – 34.99** | 6 (13%) | 5 (10.9%) | 2 (4.4%) | 3 (6.8%) | 7 (16.3%) | 11 (25%) |
| **25 – 29.99** | 13 (28.3%) | 12 (26.1%) | 19 (42.2%) | 19 (43.2%) | 3 (7%) | 1 (2.3%) |
| **20 – 24.99** | 13 (28.3%) | 12 (26.1%) | 8 (17.8%) | 11 (25%) | **0** (0%) | 1 (2.3%) |
| **15 – 19.99** | 9 (19.6%) | 11 (23.9%) | 4 (8.9%) | 4 (9.1%) | **0** (0%) | **0** (0%) |
| **< 15** | 4 (8.7%) | 6 (13%) | 1 (2.2%) | 1 (2.3%) | **0** (0%) | **0** (0%) |
| ***Total*** | *46* | *46* | *45* | *44* | ***43*** | ***44*** |
| **P-value** | **P – Value: 0.37** | | *** P-value = 0.047** | | **P-value = 0.22** | |
